# Heading for personalized rTMS in tinnitus: reliability of individualized stimulation protocols in behavioral and electrophysiological responses

**DOI:** 10.1101/2021.01.11.21249600

**Authors:** Stefan Schoisswohl, Berthold Langguth, Tobias Hebel, Mohamed A. Abdelnaim, Gregor Volberg, Martin Schecklmann

**Affiliations:** Department of Psychiatry and Psychotherapy, University of Regensburg, Regensburg, Germany; Institute of Psychology, University of Regensburg, Regensburg, Germany

**Keywords:** repetitive transcranial magnetic stimulation, tinnitus, neuronavigation, electroencephalography, reliability, rTMS personalization

## Abstract

**Background:** Repetitive transcranial magnetic stimulation (rTMS) is a non-invasive brain stimulation tool potentially modulating pathological brain activity. Its clinical effectiveness is hampered by varying results and characterized by inter-individual variability in treatment responses. RTMS individualization might constitute a useful strategy to overcome this variability. A precondition for this approach would be that repeatedly applied protocols result in reliable effects. The condition tinnitus provides the advantage of immediate behavioral consequences (tinnitus loudness changes) after interventions and thus offers an excellent model to exemplify TMS personalization.

**Objective:** The aim was to investigate the test-retest reliability of short rTMS stimulations in modifying tinnitus loudness and oscillatory brain activity as well as to examine the feasibility of rTMS individualization in tinnitus.

**Methods:** Three short verum (1Hz, 10Hz, 20Hz; 200 pulses) and one sham (0.1Hz; 20 pulses) rTMS protocol were administered on two different days in 22 tinnitus patients. Before and after each protocol, oscillatory brain activity was recorded with electroencephalography (EEG) together with behavioral tinnitus loudness ratings. RTMS individualization was executed by dint of behavioral and electrophysiological responses. Stimulation responders were identified via consistent sham-superior increases in tinnitus loudness (behavioral responders) and alpha power increases or gamma power decreases (alpha responders/ gamma responders) in accordance to the prevalent neurophysiological models for tinnitus.

**Results:** It was feasible to identify individualized rTMS protocols featuring reliable tinnitus loudness changes (55% behavioral responder), alpha increases (91% alpha responder) and gamma decreases (100% gamma responder) respectively. Alpha responses primary occurred over parieto-occipital areas, whereas gamma responses mainly appeared over frontal regions. Contrary, test-retest correlation analyses per protocol on a group-level were not significant neither for behavioral nor for electrophysiological effects. No associations between behavioral and EEG responses were given.

**Conclusion:** RTMS individualization via behavioral and electrophysiological data in tinnitus can be considered as a feasible approach to overcome low reliability on group-level. The present results open the discussion favoring personalization utilizing neurophysiological markers rather than behavioral responses. These insights are not only useful for the rTMS treatment of tinnitus but also for neuromodulation interventions in other pathologies as our results suggest that the individualization of stimulation protocols is feasible despite absent group-level reliability.

## 1. Introduction

With a prevalence of about 10% of the population, tinnitus is a rather common condition. It is characterized by the perception of ringing or hissing in the absence of a corresponding external stimulus. The majority of tinnitus cases are caused by cochlear damage leading to hearing loss [1–3], whereby the absence of auditory input from the periphery provokes maladaptive neural changes which ultimately manifest in hyperactivity of auditory and non-auditory cortical regions. These pathological alterations of the central nervous system are assumed to underly the perception of tinnitus [4–8].

Recent neuromodulation techniques attempt to reverse or modulate these pathological alterations (for a recent overview see Langguth et al. (2020) [9]. One of these is repetitive transcranial magnetic stimulation (rTMS), a non-invasive brain stimulation tool with the capability to evoke neuroplastic changes by a rhythmic administration of brief electromagnetic pulses [10–12]. By means of low frequency rTMS it is possible to inhibit cortical excitability whereas high frequency protocols are assumed to increase cortical excitability. These statements should be seen as a rule of thumb as several findings show high inter- and intra-individual variability of rTMS with a lot of subjects not behaving accordingly [13–16].

On this account, temporal 1Hz rTMS was applied in order to inhibit tinnitus-associated hyperactivity of the auditory cortex and consequently suppress the tinnitus percept [17,18].

For many years various treatment trials deployed left temporal low frequency rTMS showing greatly varying findings and high inter-individual variability in responses (for an overview see Schoisswohl et al., 2019 [19]). Given this backdrop, additional studies also examined the consequences of other rTMS approaches such as multi-site stimulation protocols [20,21], prefrontal cortex stimulation [22–24] or high frequency rTMS [25–27] as well as continuous theta-burst stimulations [25,28,29] as a treatment option for tinnitus. Nevertheless, its clinical effectiveness is still not obvious. Recent reviews and meta-analyses conclude that there is no clear indication for an effect [9,30,31], while others report potential effectiveness for its clinical administration [32,33], especially over the auditory cortex [34]. This lack of evidence may be explained by the fact that there is still a certain ambiguity concerning the correct rTMS parameters (e.g., frequency or position) for the application in tinnitus. Admittedly, the effects of non-invasive brain stimulation methods in general are subject to rigorous intra- and inter-individual variability, conceivably caused by a complex interplay of technical and physiological parameters [35–38].

A personalization of stimulation protocols might provide remedy and further enhance efficacy. Individualization of magnetic stimulation might be capable of resolving the issue of effect variability within the same type of pathology. Its implementation in neuropsychiatric research is highly dependent on phenotypes as well as the identification of appropriate and valid clinical or physiological outcome measures [39,40]. However, tinnitus provides the tremendous benefit of direct reactions from the patients’ side by virtue of changes in tinnitus perception and can therefore function as a kind of role model in rTMS personalization.

Test sessions (i.e., short stimulations with recording of immediate responses) and an administration of different protocols offer the opportunity to examine the ability of the different protocols to suppress tinnitus loudness in an individual. In several studies it has been demonstrated that tinnitus loudness can be suppressed by means of single rTMS sessions for some minutes [41–45], and that suppression patterns vary among patients [46]. The concept of rTMS personalization in the clinical context of tinnitus was first introduced by Kreuzer et al. (2017) [47]. By dint of rTMS test sessions with different types of frequencies and positions, individualized protocols for brief tinnitus suppression were identified and further used in the course of a daily treatment. These findings emphasize the feasibility of rTMS individualization and imply superiority over a standard treatment protocol by means of a higher number of treatment responders, even if no statistical differences between the standard and the individualized treatment were detected in this relatively small pilot study. In another preliminary work from our group using an e-field guided system we were able to demonstrate reliable tinnitus loudness decreases for certain stimulation parameters as well as the possibility to individualize rTMS characterized by a consistent and sham-controlled tinnitus loudness suppression in five out of five individuals [48]. Nonetheless, behavioral responses are governed by certain limitations such as their highly subjective matter and the difficulty to accurately evaluate minor changes in the tinnitus percept in many cases, which might constitute an additional reason for variability in responses.

It is widely established that tinnitus is accompanied by pathologically altered oscillatory brain activity patterns. According to the “Thalamo-cortical Dysrhythmia Model” (TCD), the assumed mechanism behind these changes are peripheral deafferentation causing thalamic low frequency activity which in turn provokes a boost in neural synchrony and increased gamma activity in auditory regions of the cortex [49–51]. An expansion of the TCD model termed “Synchronization-by-Loss-of-Inhibition-Model” (SLIM), presumes that an ancillary suppression of neurons relevant for inhibition additionally causes diminished activity in the alpha frequency range. Moreover pathological gamma in tinnitus is prompted by this alpha decrease and the concomitant loss of inhibition [52].

Several neurophysiological investigations using electro- or magnetoencephalographic measurements revealed decreased activity in the alpha band and increased activity in the delta and gamma frequency ranges [53–58]. In particular, gamma is suggested to be closely related to the actual tinnitus perception [57,59,60]. These altered spontaneous brain activities might provide a potential indicator to examine the effect of rTMS in tinnitus.

TMS-EEG investigations have already demonstrated that rTMS successfully modulates evoked and ongoing brain activity with effects exceeding the stimulation offset (neuroplastic consequences) [61–63]. In the clinical context, TMS-EEG approaches are already used for several neuropsychiatric conditions such as schizophrenia, depression or Alzheimer’s disease in order to identify and investigate the relevance of neurophysiological markers as well as to provide a more profound insight into the neuroplastic consequences of rTMS [64,65]. In case of tinnitus, a combination of rTMS and EEG might not only help to improve our understanding of the pathophysiology and altered ongoing brain activity patterns behind tinnitus but also shed light on neurophysiological markers for tinnitus suppression. As of yet, only a handful of studies investigated the consequences of single rTMS sessions on spontaneous brain activity in tinnitus. Müller et al. (2013) [46] applied different stimulation types (1 Hz, continuous theta burst, intermittent theta burst and individual alpha frequency rTMS) over the temporal cortex and aimed to detect the most efficient protocol for transient tinnitus suppression per subject. By using magnetencephalography (MEG) they could detect a significant increase of alpha power (8 – 12 Hz) in the auditory cortex associated with tinnitus suppression. Due to the absence of clear effects on a whole-group level, the authors emphasize the relevance of analysis on an individual subject level to unveil the key mechanisms behind tinnitus suppression. In this context especially modulations in the alpha frequency band might provide a potential marker for future attempts in individualizing rTMS. Schecklmann et al. [66] compared the neuroplastic consequences of rTMS in tinnitus patients with healthy controls and observed rTMS-specific modulations almost entirely for the group of tinnitus patients. A left temporal 1 Hz stimulation with 200 pulses decreased power spectra in the delta (2 – 3.5 Hz) and theta (4 – 7.5 Hz) frequency ranges as well as increased beta2 (18.5 – 21 Hz) frequency band power over frontal areas in tinnitus patients. Right prefrontal cortex 1 Hz stimulation reduced beta3 (21.5 – 30 Hz) and gamma (30.5 – 44 Hz) band activity in tinnitus patients in temporal areas. In contrast the control group exhibited enhanced beta3 and gamma power. Results emphasize the ability to modulate tinnitus-related ongoing brain activity or rather to induce neuroplastic changes in tinnitus by rTMS in accordance with the TCD model. However, on a behavioral level reliable sham-controlled decreases in tinnitus loudness were solely present in two out of 20 tinnitus patients (one after right prefrontal, one after left temporal rTMS).

Considering past findings, in particular the ability of certain rTMS protocols to transiently decrease tinnitus loudness plus modulate tinnitus-associated oscillatory brain activity, up to now there is no convincing data on test-retest reliability of the effects of single session rTMS. A test-retest design has the potential to yield essential details about effect-consistency, more specifically if rTMS produces reliable and valid modulations. Findings may promote decisions in choosing appropriate rTMS protocols for daily treatments. Due to the overall high variability of non-invasive brain stimulation effects, it is of utmost importance to identify reliable brain stimulation techniques to cope for this variability. Hence, one of the objectives of the current experiment was to scrutinize the test-retest reliability of different rTMS protocols in short-term tinnitus suppression and modifying resting state EEG activity. For this purpose the study design used in Schoisswohl et al. (2020) [48] with pre-post rTMS tinnitus loudness ratings was expanded by electrophysiological measures before and after each rTMS. Furthermore, it was reduced by the factor stimulation position since the majority of patients (three out of five) responded to a stimulation over temporo-parietal regions. An e-field guided stimulation over the left and right temporo-parietal junction allows for a more precise investigation of the parameter frequency, since past studies show inconsistent results in this regard.

Beyond that, we wanted to undertake further research on the personalization of rTMS by means of the model tinnitus. The aim was to assess the feasibility of identifying an rTMS protocol for consistent and sham-superior brief tinnitus suppression per patient (behavioral response). In light of previous work from Müller et al. (2013) [46] together with the current prevalent neurophysiological models in tinnitus, we wanted to go one step beyond and strive for a rTMS personalization based on electrophysiological responses. The specific objective was to ascertain a protocol per patient, which is able to produce consistent and sham-superior increases in alpha or decreases in the gamma frequency band (EEG response) respectively. Based on past research, we hypothesize that for the majority of subjects it is feasible to identify a personalized rTMS protocol by means of behavioral or EEG responses.

Additionally, we were interested if an EEG response in the alpha or gamma band is related to a behavioral response and vice versa.

## 2. Material and Methods

### 2.1 Participants

N = 22 patients (5 female) with chronic subjective tinnitus, recruited from the Interdisciplinary Tinnitus Centre in Regensburg, Germany, participated in this experiment. Participants had to comply to the following inclusion criteria: age between 18 and 75 years; German-speaking; no or stable treatment with psychoactive drugs; absence of severe somatic, neurological or psychiatric conditions (e.g., acute psychosis, severe depression, alcohol and/ or substance abuse or known brain tumor); no simultaneous participation in other tinnitus-related experiments or treatments. Further, participants had to exhibit no contraindications with respect to TMS (e.g., epilepsy or state after traumatic brain injuries; cf. [67,68]) as well as magnetic resonance imaging (MRI) (e.g., heart pacemaker, vascular clamp, implanted insulin pump or severe claustrophobia). Each participant received detailed clarification about study aim, procedure and applied methods as well as potential adverse effects related to TMS and gave written informed consent before study start. The study was approved by the ethics committee of the University of Regensburg, Germany (ethical approval number: 17-820-101).

### 2.2 Test session procedure

Ahead of the rTMS tests sessions, participants’ hearing thresholds for the frequency range of 125 Hz to 8 kHz were determined by the use of a clinical audiometer (Madsen Midimate, 622D; GN Otometrics, Denmark). In addition, structural MRI brain scans (T1) were undertaken with a MAGNETOM 1.5 Tesla scanner (Siemens, Germany) for neuronavigated rTMS and participants were requested to complete German versions of the Tinnitus Handicap Inventory (THI; [69,70]) the Tinnitus Sample Case History Questionnaire (TSCHQ; [71]) and the European School for Interdisciplinary Tinnitus Research Screening Questionnaire (ESIT-SQ; [72]).

The actual rTMS test sessions lasted approximately three hours and took place on two different days with a maximum inter-session interval of two days, whereby daytime was always the same (± 1 hour). Both test session days followed the same procedure. On each day, subjects were stimulated with four different rTMS protocols over the left temporo-parietal junction (TPJ) and with the same four protocols over the right TPJ (see section repetitive transcranial magnetic stimulation). Thus, a total number of 8 different rTMS protocols was applied on each test session day. Magnetic stimulation protocols were administered in a randomized sequence on each day with the exception that the order of hemisphere was inverted (if the stimulation sequence on the first day started over the left hemisphere, it began with a stimulation over the right hemisphere on the second day). During three minutes before and after each stimulation patients were requested to focus on their tinnitus and verbally rate the current loudness of their tinnitus perception on a numeric rating scale ranging from 0% to 110% at seven different points in time with an inter-rating interval of 30 seconds (T0, T30, T60, T90, T120, T150 and T180). The numeric rating scale was anchored at 0% (absence of tinnitus) and at 110% (tinnitus loudness increase of 10%), whereby 100% corresponds to the patients usual perceived tinnitus loudness. Concurrent to these ratings, resting state oscillatory brain activity was recorded for three minutes each by means of EEG. For this purpose, participants were instructed to sit calmly, focus on a certain point in the room and avoid unnecessary eye blinks or muscle movements. Subsequent to each post stimulation loudness rating/ EEG recording, participants had to evaluate the level of discomfort caused by the applied rTMS protocol on a numeric rating scale from 0 to 10, whereas 10 corresponds to utmost discomfort or rather intolerability. **Figure 1** provides an illustration of the entire test session proceeding.

**Figure 1.**
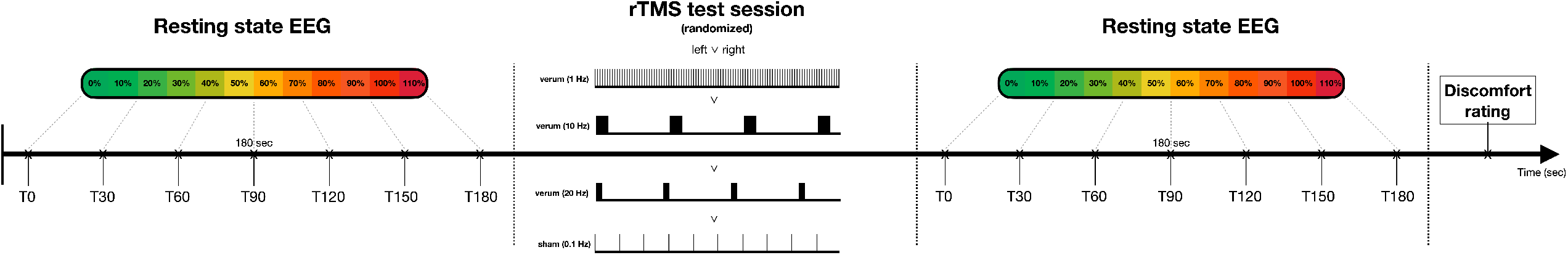
Test session procedure. Test sessions were conducted on two different days according to the same procedure. Three different verum rTMS protocols (1 Hz, 10 Hz, 20 Hz) with 200 pulses each and one sham protocol (0.1 Hz) with 20 pulses were administered over the left and right temporo-parietal junction in a randomized order. Thus, a total number of 8 different rTMS protocols was applied on each test session day. Before and after each magnetic stimulation, participants were requested to rate the subjective loudness of their tinnitus on a numeric rating scale from 0% to 110% (0% - complete absence of tinnitus; 100% - no change; 110% - 10% loudness increase) at seven points in time (30 seconds inter-rating interval). In parallel to these ratings, resting state brain activity was recorded with electroencephalography (EEG) for three minutes. At the end of each rTMS protocol (pre-rating/ recording – stimulation – post-rating/ recording), patients had to rate the induced level of discomfort on a numeric rating scale from 0 to 10 (10 – utmost discomfort).

### 2.3. Repetitive transcranial magnetic stimulation (rTMS)

An e-field guided rTMS system (NBT System 2; Nexstim Plc., Finland) together with individual structural MRI scans (T1) was utilized for the test sessions. Prior to the first session, participants’ resting motor threshold (RMT) was determined according to the same procedure as already described in Schoisswohl et al. (2020) [48]. Thereby, various positions over the left primary motor cortex were stimulated until several motor evoked potentials (MEP) derived from the thenar muscles of the right hand with a peak-to-peak amplitude of > 50 μV were visible. The stimulation position with the highest MEP amplitude was repeated via the neuronavigation system and single pulses with intensity shifts were applied. The lowest stimulation intensity which produced MEPs with an amplitude of at least 50 μV in half of the administered pulses was defined as subjects’ RMT. Additionally, the RMT determination was repeated with a mounted EEG cap using the same motor hotspot. Magnetic stimulation consisted of 1 Hz, 10 Hz and 20 Hz protocols with 200 pulses each at 110% RMT, whereas a series of 20 pulses at 0.1 Hz was deployed as an active control condition (sham) with no assumed neuroplastic consequences [73,74]. Each protocol was administered over the left and right TPJ, resulting in a total number of eight different rTMS protocols with approximately six minutes of inter-protocol intervals (3 minutes of pre and post rTMS EEG recordings; if applicable – a waiting period until subject’
ss tinnitus loudness returned to the initial level). An uncooled coil was used for the test sessions, as the cooled coil produces additional noise and we aimed at minimizing the stimulation noise, as it could confound both behavioral and neurophysiological effects. Stimulation positions were localized according to the electrode positions CP5 and CP6 (10-20 System). By virtue of the application of an e-field guided neuronavigation system and the opportunity for online visualization of strength and direction of the TMS-induced e-field on subjects’ anatomical scans, each stimulation was administered with the induced e-field perpendicular to the sulcus of the brain area of interest. In order to ensure that the stimulation positions respectively the induced e-field remained the same over the two test sessions, both electrode positions were tagged with a digitization pen and marked on the individual anatomical images. Next, the coil was placed accordingly, a single pulse with an intensity of 10% RMT was given and the corresponding position of the coil was saved in the neuronavigation system. With the aid of a system-intern aiming tool, the exact position of the coil could be repeated with respect to centering, rotation and tilting. After each protocol the position was verified and adjusted if necessary. For safety reasons all participants were wearing in-ear plugs during the whole stimulation procedure.

### 2.4. Electroencephalography

#### 2.4.1. Data acquisition and preprocessing

Participants’ electrophysiological resting state brain activity was recorded for three minutes before and after each of the eight rTMS protocols by means of EEG with a BrainAmp DC system (Brain Products GmbH, Germany) in combination with an Easycap elastic electrode cap (Easycap GmbH, Germany) consisting of 64 electrodes placed according to the 10-20 System (GND: AFz) and the software Brain Vision Recorder 1.20 (Brain Products GmbH, Germany). EEG measurements were online referenced to FCz and recorded at a sampling rate of 500 Hz. Impedances were kept lower than 10 kΩ during the whole recording.

Preprocessing of raw EEG data was conducted with a custom-built pipeline in Matlab (Matlab R2018b; Mathworks, USA) by employing functions implemented in the EEGLAB toolbox [75]. In a first step, EEG data was filtered between 1 Hz and 45 Hz and segmented into 2 second epochs. Afterwards the data was visually inspected and single epochs featuring artifacts or verbal tinnitus loudness ratings were removed from the signal. For each dataset the first and last two epochs were rejected as well, and noisy or deviant sensors were identified for subsequent interpolation. In order to identify and reject vertical and horizontal eye movements, an independent component analysis (ICA; fastICA toolbox http://research.ics.aalto.fi/ica/fastica/) was executed. In a next step, datasets were re-referenced using an average reference and the online reference electrode FCz was reconstructed and added as a data channel. Previously identified channels with high signal-to-noise ratio were interpolated using the spherical spline method [76]. As a last step, preprocessed datasets underwent a final visual inspection.

#### 2.4.2. Power spectra

Power spectra analysis was performed with the Fieldtrip toolbox [77] in Matlab. Fast Fourier Transformation (FFT, “mtmfft”) and a hanning window with 1 Hz spectral smoothing was used for the calculation of pre and post stimulation power spectra for each channel per rTMS protocol. Obtained power spectra underwent a normalization through a division of the spectral power of every single frequency by the average spectral power of the whole frequency spectrum. Grand averages were calculated for every pre and post stimulation dataset per channel and the following frequency bands: delta (2 – 3.5 Hz), theta (4 – 7.5 Hz), alpha (8 – 12.5 Hz), beta (13 – 32.5 Hz) and gamma (33 – 44 Hz) [66,78]. RTMS protocol specific pre-to-post power changes in the respective frequency bands were obtained by subtracting the pre stimulation from the post stimulation power for each channel (Δ power = post – pre). Channels Iz, TP9 and TP10 which usually contain noisy signal were excluded, resulting in 60 channels for further statistical analysis.

### 2.5. Statistical analysis

All subsequent statistical analyses were performed with the statistics software R (R version 4.0.3; R Foundation for Statistical Analysis, Austria) using the packages “psych” and “ggstatsplot”. Assumptions for parametric testing were examined with Shapiro-Wilk-Tests (normal distribution) and F-Tests (sphericity) and if violated non-parametric tests were performed. Significance levels were set to ≤ .05 for all analysis, whereby < .10 was considered as a statistical trend. Differences between left and right hearing level, RMT as well as induced e-field strength with and without mounted EEG cap were analyzed with paired sample t-tests and Wilcoxon signed-rank tests respectively.

#### 2.5.1. Reliability analysis of TMS-induced tinnitus loudness changes and discomfort

Pre stimulation ratings were averaged (T0 – T180) to receive a stable tinnitus loudness baseline for each protocol per subject. TMS-induced tinnitus loudness changes were calculated via a subtraction of the mean baseline loudness level from the mean post stimulation loudness level (Δ loudness = post – baseline). Test-retest reliability of TMS-induced mean tinnitus suppression as well as discomfort evaluation over both test session days was analyzed by means of Spearman correlations for each protocol. P-values were adjusted for multiple comparisons using the Holm-Bonferroni method [79].

#### 2.5.2. Reliability analysis of TMS-induced changes in oscillatory brain activity

Test-retest reliability of protocol-specific pre-post power spectra changes were analyzed with Spearman correlations. Therefore, the obtained changes were correlated between the first and second test session on a single sensor level per rTMS protocol and frequency band. Thus, 300 correlations were calculated per rTMS protocol (5 frequency bands x 60 sensors). Obtained p-values of sensor correlations were adjusted according to the number of sensors separated per frequency band and protocol using the Holm-Bonferroni method [79]. In case of significant results, p-values were further corrected for the number of applied protocols akin to an adjustment for multiple comparisons of behavioral results. We were exclusively interested in positive correlation outcomes, representing reliability of TMS effects. Significant positive correlations of channel pairs were identified via correlation matrices and projected on a 60-channel topographical scalp map applying a color-coding for received correlation coefficients (r_s_). A minimum number of two neighboring electrodes featuring significant positive correlations (electrode cluster) was considered as a reliable TMS-induced change in power spectra. Triangulation of 2d sensors was utilized for neighboring sensor identification.

#### 2.5.3. rTMS individualization via responder identification using behavioral and electrophysiological data

Additional to test-retest reliability analysis, rTMS individualization was examined based on single subject responses concerning loudness evaluation and electrophysiological consequences. Behavioral response to one of the verum interventions was defined as a mean tinnitus loudness suppression (compare section Reliability analysis of TMS-induced tinnitus loudness changes and discomfort evaluation) superior to sham stimulation (Δ loudness verum > Δ loudness sham) on both test session days for the same rTMS protocol and stimulated hemisphere. EEG responders were defined for the alpha (8 – 12.5 Hz) and gamma (33 – 44 Hz) frequency bands separately. In particular we were interested in alpha power increases and gamma power decreases from pre to post stimulation. As indicated in the introduction, alpha increases and gamma decreases are the most valid biomarkers for tinnitus reductions as indicated by literature review and expert knowledge. For each subject and protocol, the top 20% of channels exhibiting alpha increases/ gamma decreases were detected and scrutinized with regards to sham-superiority per day.

If a sham-superior alpha increase respectively gamma decrease was observed in the same sensor on both days plus the corresponding pattern occurred in a minimum number of two adjacent channels, the subject was designated as either alpha or gamma responder. Associations of a behavioral response to any of the eight protocols with an electrophysiological response in the alpha or gamma frequency band and vice versa were examined with Fisher’
ss exact tests due to cell frequencies less than five.

## 3. Results

### 3.1. Sample characteristics

Participants were aged from 43 to 69 years (M = 57.05, SD = 6.79) and had an average tinnitus duration of 131.64 months (SD = 116.76). The majority reported a bilateral tinnitus perception as well as loudness fluctuations. No difference in hearing loss between the left and the right ear was observed, t_(13)_ = -1.36, p = .198. THI severity grades ranged from mild (grade 2) to catastrophic (grade 5) and manifested on average in severe severity (grade 4) for the whole sample (M = 58.49, SD = 19.55). As expected, the RMTs were significantly higher with a mounted EEG cap, p < .001. Unexpectedly, the corresponding strength of the induced e-field was similarly higher with an EEG-cap, t_(21)_ = -6.07, p < .001. Although the motor hotspot for RMT determination was identical as well as coil centering and rotation, differences in the e-field might derive from difficulties in the adjustment of coil tilting due to distances to the scalp as well as the concomitant stimulation at higher intensities.

Information about participant characteristics can be seen from **table 1**. Besides anticipated side effects such as discomfort during rTMS (in particular by rapid muscle contraction caused by high frequency protocols) or temporary tinnitus loudness increases after stimulation, no additional side effects could be observed in the current sample. Spearman correlations showed significant positive correlations for discomfort evaluations of each applied rTMS protocol between the two test sessions (cf. **figure S1** in the supplemental material).

**Table 1.** Participants characteristics

| N (female) | 22 (5) |  |  |  |
| --- | --- | --- | --- | --- |
| Handedness (left/ right/ both) (4 missings) | 0/ 14/ 4 |  |  |  |
| Tinnitus laterality (left/ right/ both/ inside head) (4 missings) | 1/ 1/ 14/ 2 |  |  |  |
| Tinnitus loudness fluctuation (yes/ no) (4 missings) | 14/ 4 |  |  |  |
|  | **M ± SD** | **Md** | **Min** | **Max** |
| Age (years) | 57.05 ± 6.79 | 57.50 | 43.00 | 69.00 |
| Tinnitus duration (months) (2 missings) | 131.65 ± 116.79 | 102.00 | 5.00 | 420.00 |
| Hearing loss left (dB) (8 missings) | 23.42 ± 9.72 | 22.22 | 7.22 | 41.67 |
| Hearing loss right (dB) (8 missings) | 27.87 ± 14.29 | 23.06 | 7.78 | 61.88 |
| THI score (0-100) | 58.49 ± 19.55 | 58.00 | 30.00 | 90.00 |
| Tinnitus loudness (%) (6 missings) | 70.31 ± 19.87 | 80.00 | 30.00 | 90.00 |
| RMT (%) | 33.95 ± 5.33 | 33.00 | 25.00 | 46.00 |
| RMT (%) + EEG cap | 41.91 ± 9.65 | 41.00 | 30.00 | 66.00 |
| Electrical field (V/m) | 54.47 ± 13.90 | 52.50 | 27.00 | 86.00 |
| Electrical field (V/m) + EEG cap | 66.66 ± 15.38 | 66.25 | 44.00 | 99.00 |

M = mean; SD = standard deviation; Md = Median; Min = minimum; Max = maximum; THI = Tinnitus Handicap Inventory; RMT = resting motor threshold, EEG = Electroencephalography

### 3.2. Test-retest reliability of TMS-induced changes in tinnitus loudness and oscillatory brain activity

No significant correlations were observed for the two test session days with regards to mean tinnitus loudness changes for any of the protocols (cf. **figure 2**). It should be noted, that without considering corrections for multiple comparisons, a significant test-retest reliability was found for left hemispheric stimulation with 10 Hz and right hemispheric stimulation with 20 Hz.

**Figure 2.**
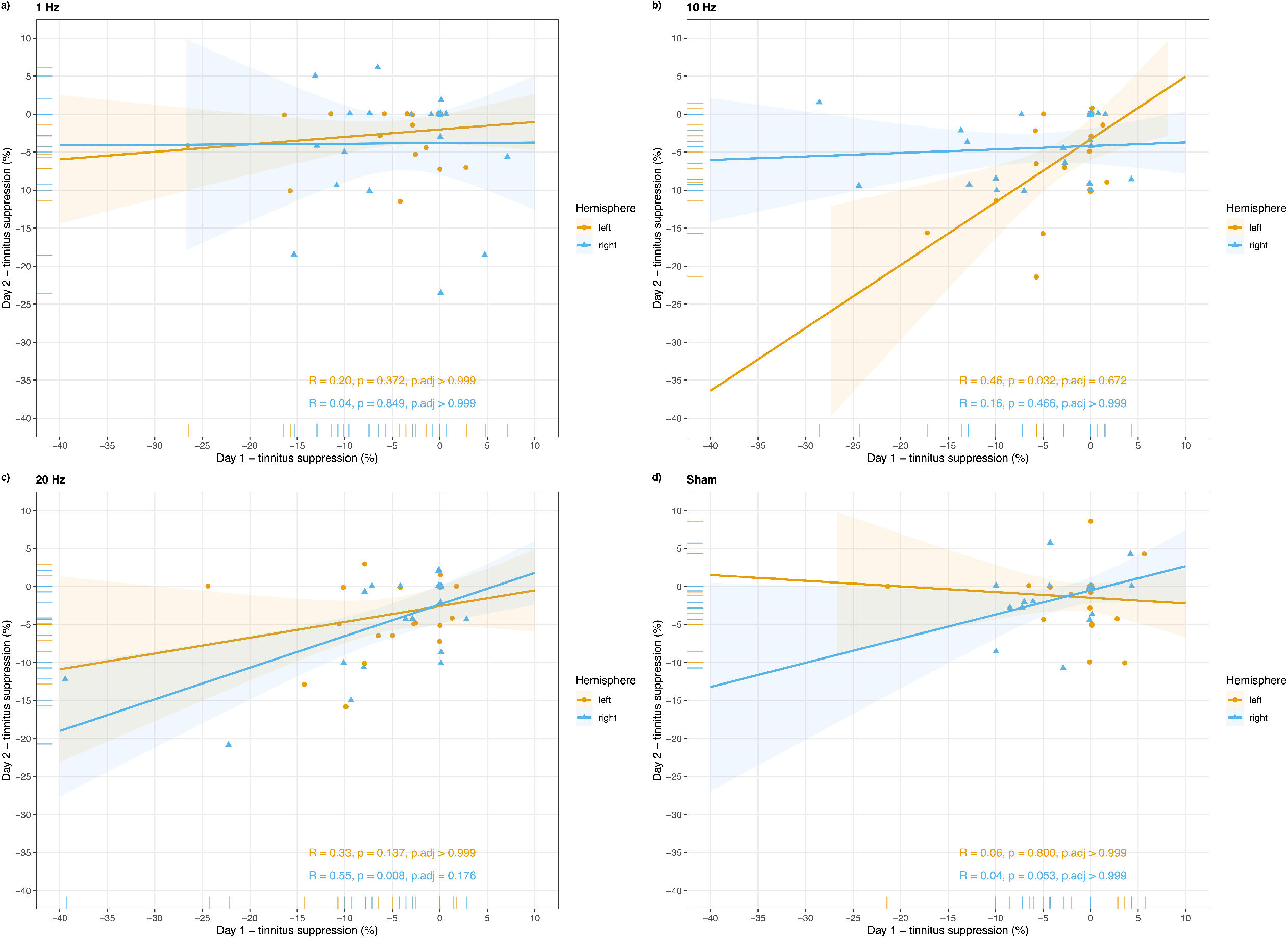
Reliability of TMS-induced changes in tinnitus loudness. Spearman correlations of rated tinnitus suppression (%) per rTMS protocol between the first and the second test session day are illustrated. Colored ribbons represent confidence intervals (95%). Reliable changes were observed for a (b) 10 Hz stimulation over the left temporo-parietal junction and a (c) 20 Hz stimulation over the right temporo-parietal junction. Both correlations do not withstand a correction for multiple comparison and should therefore not be overrated and interpreted only exploratively.

On the basis of our predefined reliability criteria, reliable changes in oscillatory brain activity were solely present for 1 Hz stimulation of the right TPJ, exclusively for the gamma frequency band. As can be seen from **figure 3**, these reliable changes were primarily observed over parieto-occipital regions, whereby neighboring channels PO7 and O1 constitute a significant reliable cluster. If corrected for multiple comparisons, this effect disappears and is therefore only conditionally interpretable.

**Figure 3.**
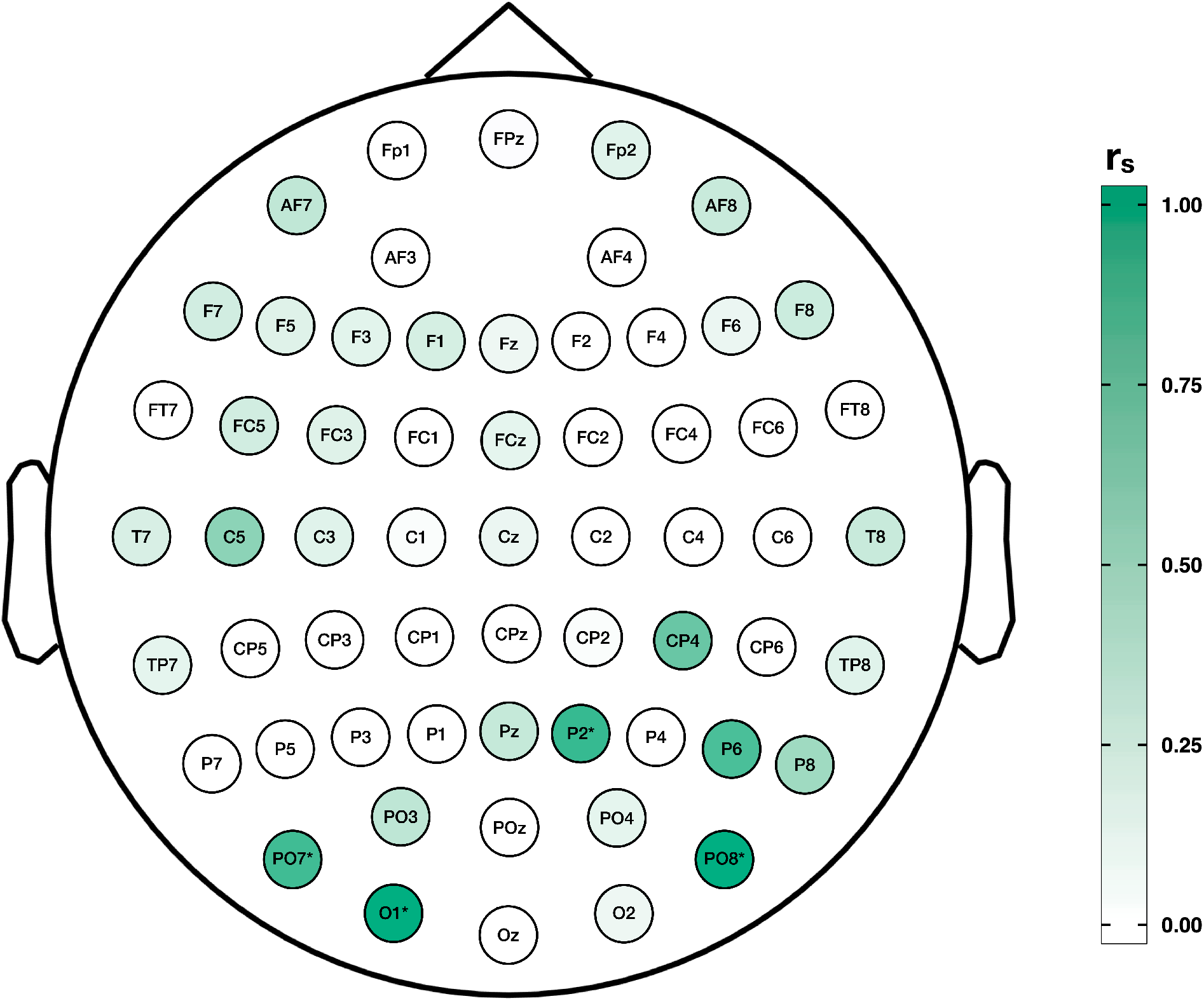
Reliability of TMS-induced changes in oscillatory brain activity. Spearman correlations coefficients (r_s_) of right hemispheric 1 Hz rTMS-induced changes in the gamma frequency band (33 – 44 Hz) for the two test session days are projected on a topographical map per electrode. Significant reliable electrodes are highlighted with asterisks. Separate reliable changes are primary present over parieto-occipital electrodes, whereby neighboring channels PO7 and O1 form a significant reliable cluster. However, this effect disappears after a correction for multiple comparison and is therefore only conditionally interpretable.

### 3.3 rTMS individualization using behavioral and electrophysiological data

In 12 out of 22 (55%) participants it was possible to identify an individualized rTMS protocol for short-term tinnitus suppression characterized by sham-superiority on both test session days. From those 12 patients 10 (83.33%) responded to more than one stimulation type. Detailed information about individual responses together with the induced mean tinnitus suppression (averaged over both test session days) can be found in **table 2**. As presented in **table 3** the majority of subjects responded to a stimulation over the left TPJ with 20 Hz (n = 7) and 10 Hz (n = 6), and over the right TPJ with 20 Hz (n = 5).

**Table 2.**
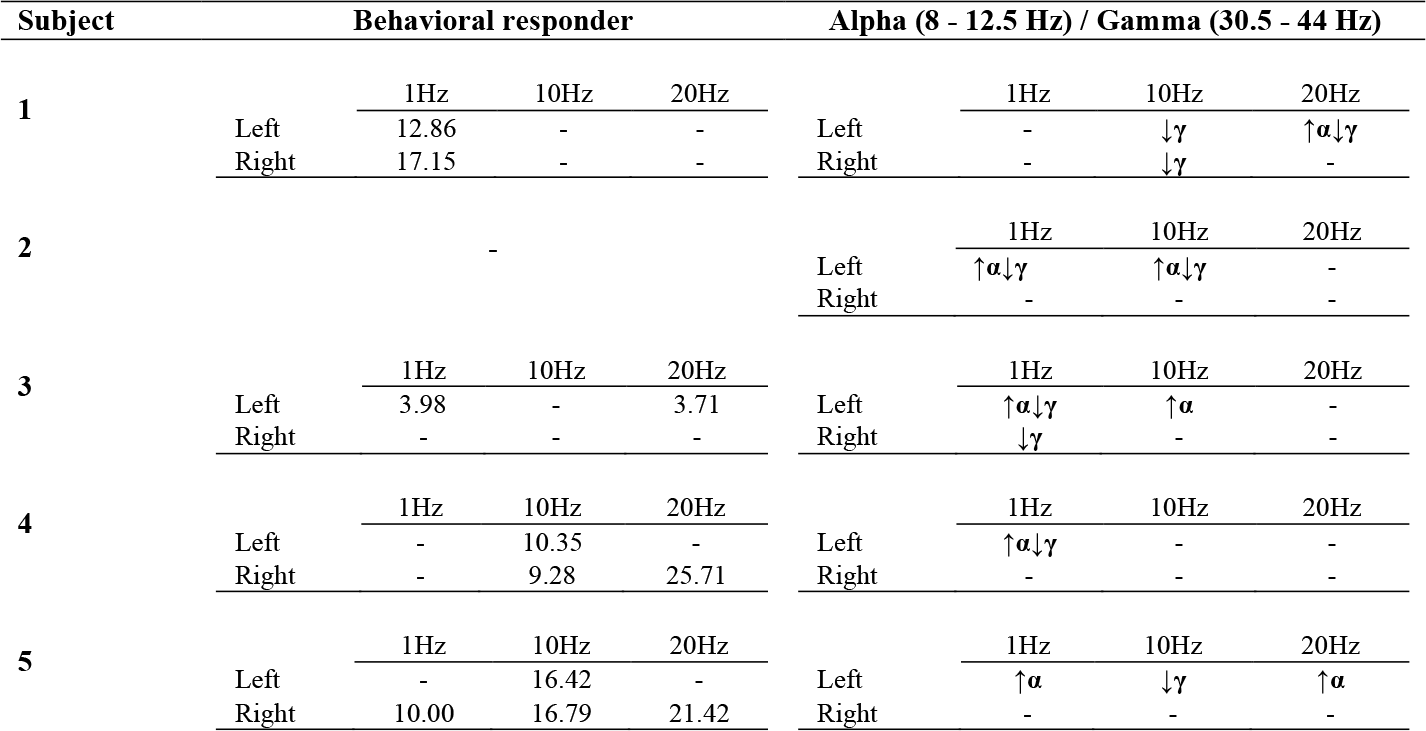
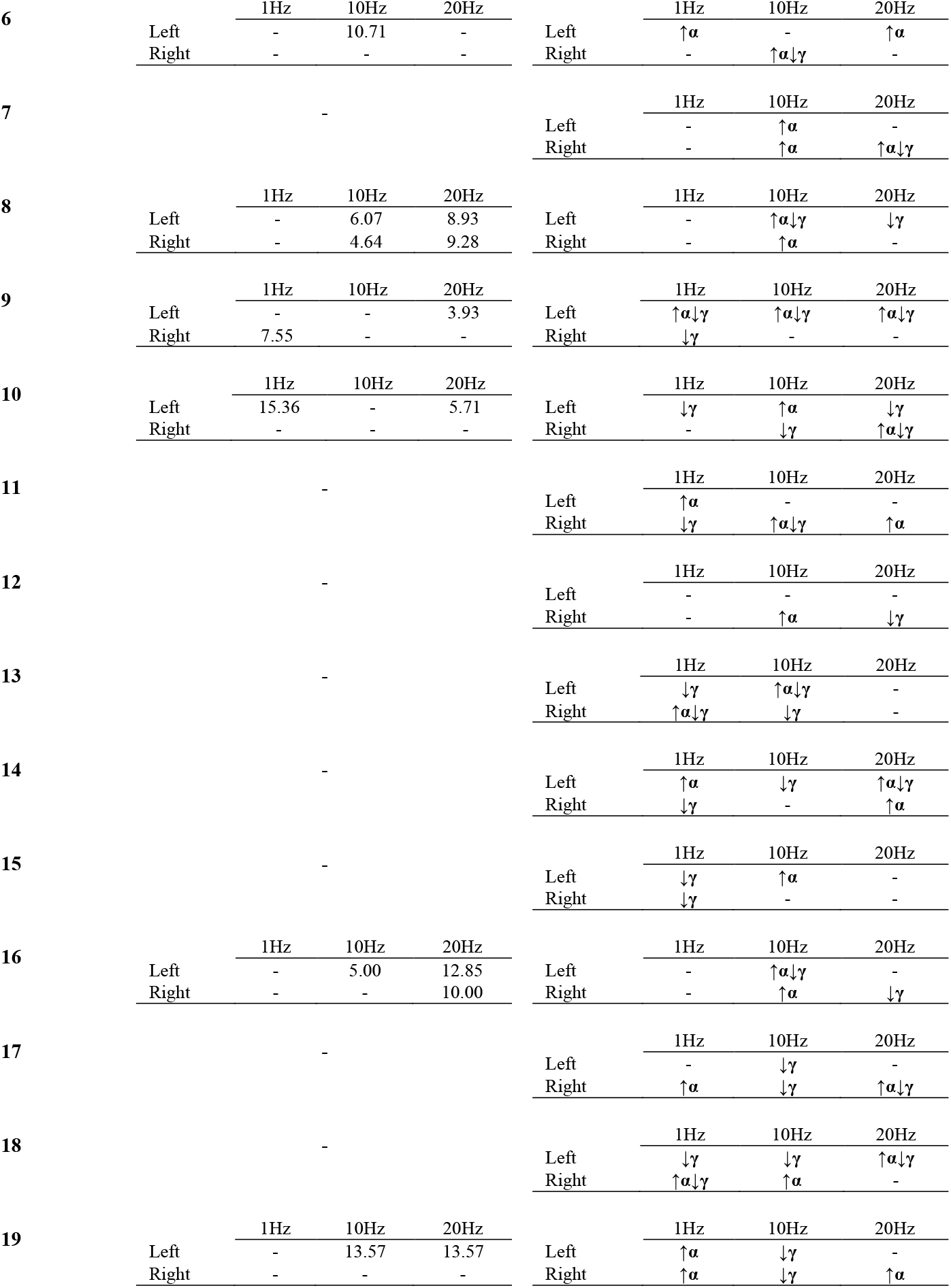
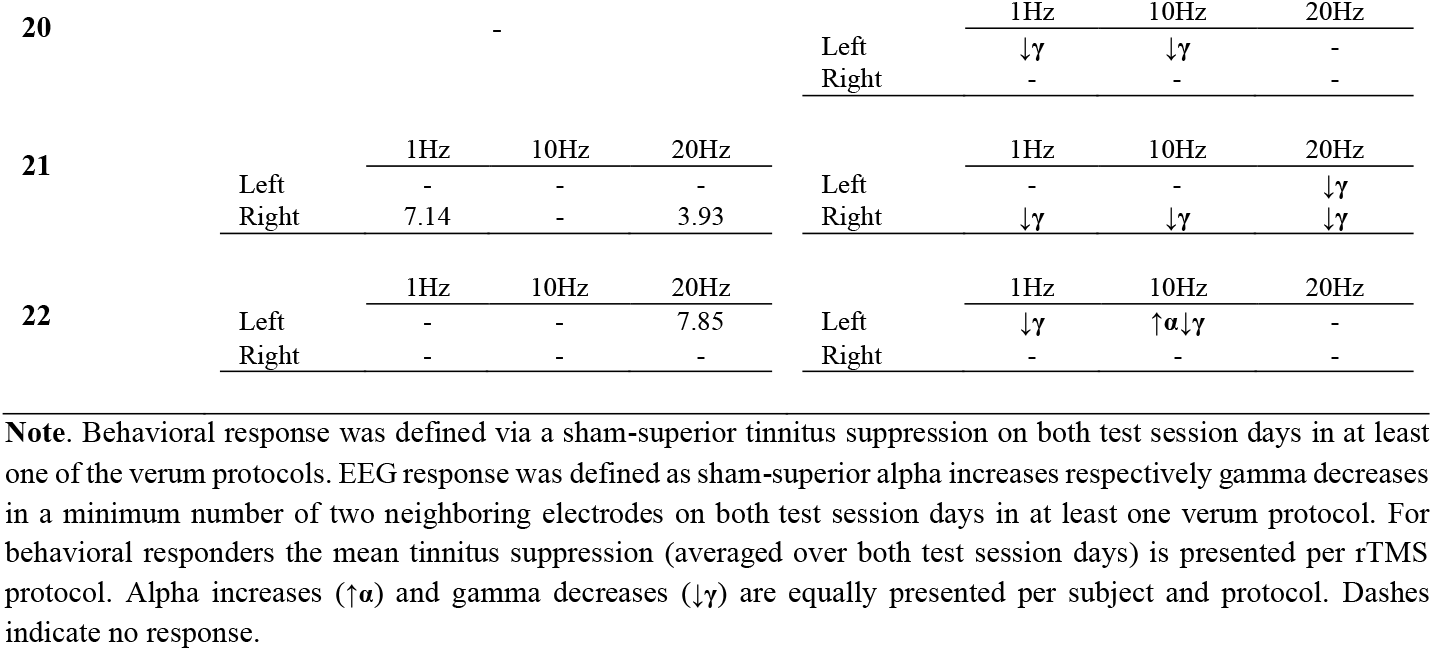
Individual test session responses per protocol – behavioral and EEG responder

**Table 3.** Summary of behavioral and electrophysiological responder per rTMS protocol

|  | Behavioral responder | ↑Alpha | ↓Gamma |
| --- | --- | --- | --- |
| Total number of responders (N) | 12 | 20 | 22 |
| Number of responders per protocol (N) |  |  |  |
| Left 1 Hz | 3 | 9 | 10 |
| Right 1 Hz | 4 | 4 | 8 |
| Left 10 Hz | 6 | 10 | 13 |
| Right 10 Hz | 3 | 7 | 8 |
| Left 20 Hz | 7 | 6 | 7 |
| Right 20 Hz | 5 | 6 | 6 |

EEG responders were defined by sham-superior alpha power increase or gamma power decrease in minimal two neighboring electrodes on both test sessions. Using this criterion 20 alpha responders (90.91%) were identified, showing alpha power increases for at least one of the six used verum protocols. Left TPJ stimulation with 10 Hz (n = 10) or 1 Hz (n = 9) and right TPJ stimulation with 10 Hz (n = 7) produced the most alpha responders (cf. **table 3**). Electrophysiological responses in the gamma band were present in each investigated subject (100%) for at least one protocol. Subjects responded mainly to left TPJ stimulation with 10 Hz (n = 13) or 1 Hz (n = 10) and right TPJ stimulation with 10 Hz (n = 8) or 1 Hz (n = 8) (cf. **table 2**). Interestingly, 75% of alpha and gamma responders showed consistent power increases/decreases for more than one protocol. **Table 3** presents an overview of alpha increases and gamma decreases per rTMS protocol on a single subject level. Fisher’
ss exact tests showed no significant association neither for behavioral responders with alpha or gamma responders nor the other way around.

Electrodes featuring sham-superior alpha/gamma modifications on both days are presented in **figure 4** and **figure 5** by means of topographical maps displaying the quantity of electrodes within alpha and gamma responders per rTMS protocol. It is apparent from **figure 4** that alpha responses primary appeared in parieto-occipital regions on both hemispheres, whereas responses in the gamma range were especially present over frontal regions bilaterally and to some extent over parieto-occipital parts as outlined in **figure 5**. Detailed electrode information per subject and rTMS protocol can be found in the supplementary material for alpha (**table S1**) and gamma responder (**table S2**).

**Figure 4.**
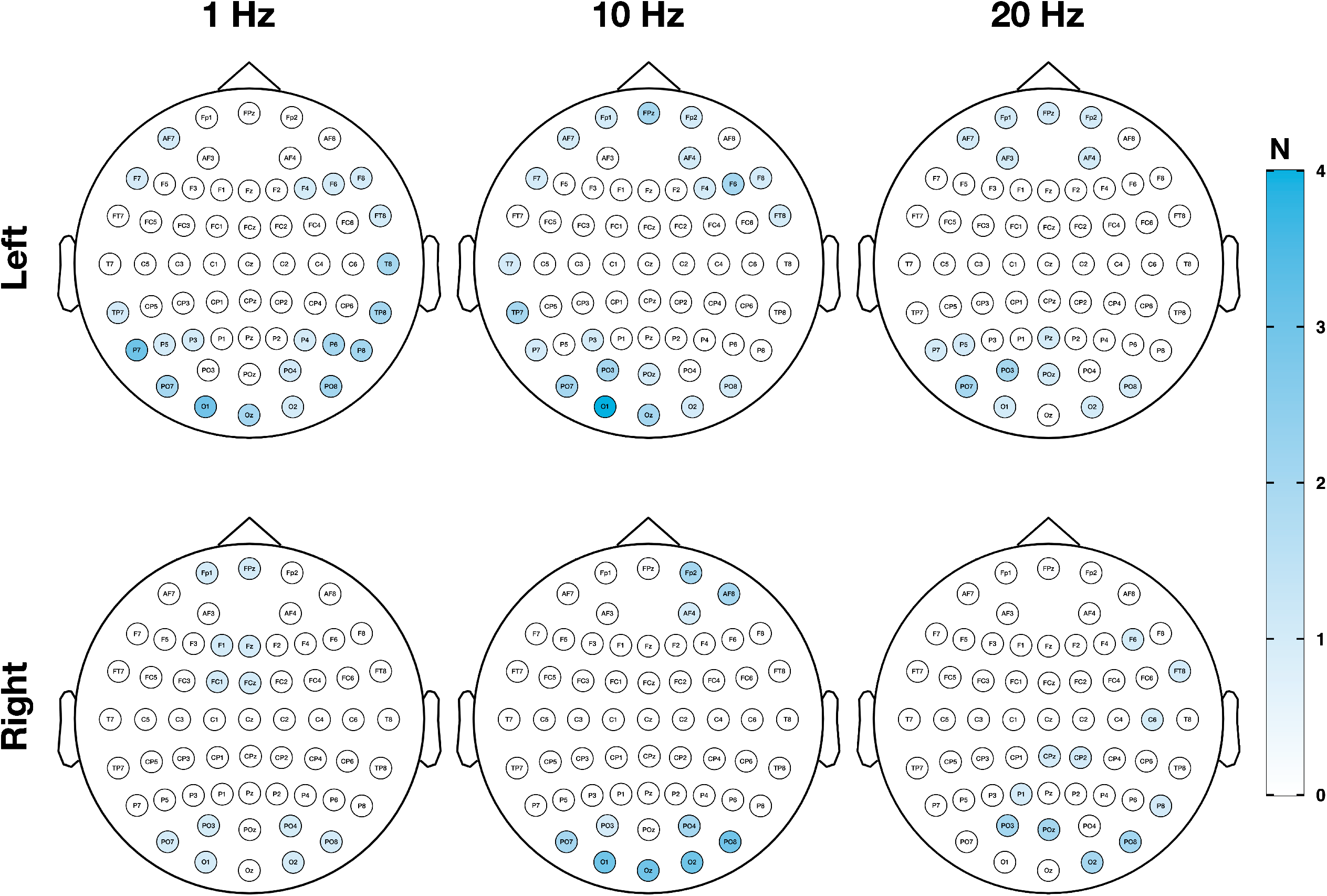
Topography of electrophysiological response – alpha responder. Topography and quantity of sensors showing sham-superior increases in the alpha frequency band in a minimum number of two neighboring channels on both test session days are illustrated per rTMS protocol. Responses in the alpha frequency band (8 – 12.5 Hz) to one of the verum protocols primarily occurred over parieto-occipital electrodes.

**Figure 5.**
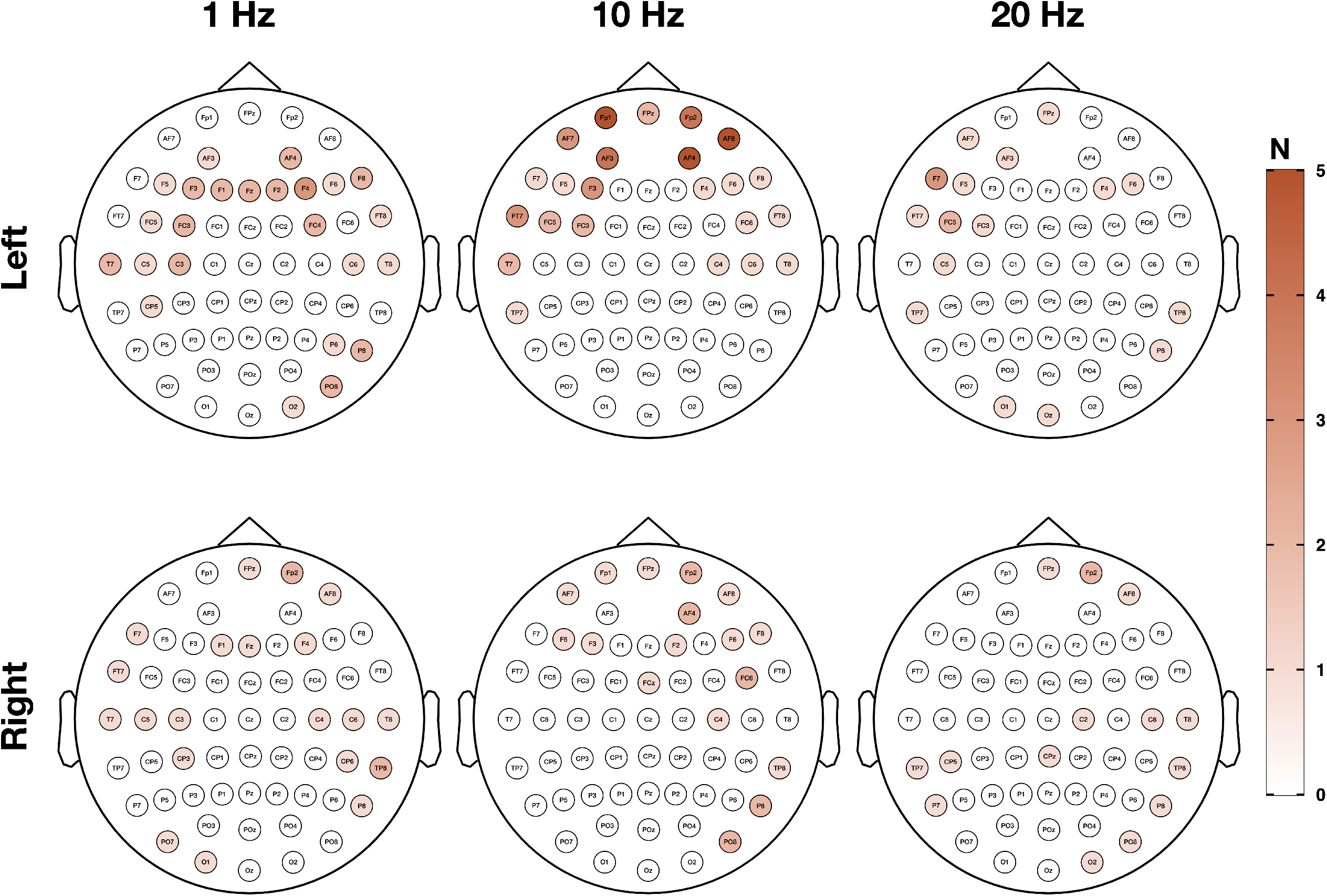
Topography of electrophysiological response – gamma responder. Topography and quantity of sensors showing sham-superior decreases in the gamma frequency band in a minimum number of two neighboring channels on both test session days are illustrated per rTMS protocol. Responses in the gamma frequency band (33 – 44 Hz) to one of the verum protocols mostly appeared over frontal electrodes on both hemispheres and partially over parieto-occipital sensors.

## 4. Discussion

The goals of this experiment were to ascertain, whether it is possible to identify treatment protocols that provide reliable effects both on the behavioral and on the electrophysiological level, as this is a precondition for individualizing treatment. We performed two test sessions at which six active and two sham protocols were administered. We evaluated reliability by comparing tinnitus suppression effects and electrophysiological effects on an individual level by responder identification. Moreover, we performed whole-group analysis in which we correlated the behavioral and electrophysiological effects of all stimulation protocols between both test sessions. The main finding of our study was, that in 55% of patients we could identify a stimulation protocol that reduced tinnitus loudness transiently in both sessions reliably more than sham stimulation. When we defined response by the effects on oscillatory brain activity, the number got even higher with 20 out of 22 patients in which a specific rTMS protocol increased alpha more than sham and with 22 out of 22 in which a specific rTMS protocol decreased gamma more than sham. These observations are in accordance with our a priori assumptions of rTMS personalization in the majority of subjects as well as findings from previous studies.

Correlation analysis for the whole group revealed reliable changes in tinnitus loudness after left TPJ stimulation with 10 Hz and right TPJ stimulation with 20 Hz. With respect to modulatory effects of rTMS on oscillatory brain activity, reliable changes could be obtained after right TPJ stimulation with 1 Hz in the gamma frequency band over parieto-occipital regions. However, these findings do not withstand a correction for multiple comparison and should therefore not be overrated and interpreted only exploratively. Nevertheless, the observed effect of rTMS on gamma activity fits well with a previous TMS-EEG study where right frontal 1 Hz rTMS decreased gamma band activity over right temporal regions in tinnitus patients, whereas it increased gamma activity in healthy controls [66]. Gamma activity is supposed to be strongly connected with tinnitus perception [57,59,60]. However, as mentioned above, the effects of the first and the second session were not significantly correlated when statistical corrections for multiple comparisons were performed.

Taking our findings together we can conclude that it is possible to identify responders for specific rTMS protocols, who demonstrate a reliable behavioral or electrophysiologic response on a specific rTMS protocol. However, if we investigate the effects of a given protocol for the whole sample, the variability of the effect is relatively large from session to session.

Presumably the effects depend on the brain state before or during rTMS [80] and this brain state might vary from session to session. In the current study the experimental design with six active and two sham protocols per session might have increased this variability, as after-effects from preceding protocols cannot be excluded as confounding factors.

The limited test-retest reliability corroborates the claim that effects of rTMS in general [37,38,81–83] but also treatment effects in tinnitus [9,84] are subject to high variability. Thus, it is most likely that single subject impacts simply do not transfer to statistical analysis on a group level, highlighting the need and importance of personalized rTMS approaches.

On the other hand, it has been demonstrated that patients, who benefit once from rTMS have a high chance to benefit from repeated rTMS treatment [85,86].

This clinical observation fits with our finding to identify treatment responders to specific rTMS protocols.

Our results corroborate also findings from several previous studies in which the feasibility of a customization of rTMS for tinnitus via test sessions has been demonstrated [47,48]. The responder rate of 55% in our study (12 out of 22) is in a similar range as in two previous studies (Kreuzer et al. (2017) - 50% responder [47]; Schoisswohl et al. (2020) - 100% responder [48]). However, both studies deployed magnetic stimulations over either prefrontal cortical regions or different positions over the superior temporal gyrus, whereas the current study targeted only a single region of the temporal cortex. The present goal was to examine the parameter frequency in more detail. Interestingly the vast majority of participants responded to high frequency stimulations over the left TPJ. It is assumed that high frequency protocols produce increases of cortical excitability [15]. Consequently, high frequency rTMS is most likely not able to reduce tinnitus-associated temporal hyperactivity. But it has been suggested, that the induced effects can vary or shift across different targets of the cortex [43] and are dependent on the intrinsic state of the brain region prior to stimulation [80,87–89]. Based on this it could be the case that pathological auditory cortex hyperactivity as an intrinsic state of the “tinnitus brain” might cause a reversal of supposed high frequency effects (shift from excitatory to inhibitory). In accordance with the present results, previous studies have demonstrated responder rates between 40% and 57% in single session experiments with high frequency stimulation protocols [41,42,45,90]. Likewise treatment studies with high frequency rTMS over the temporo-parietal cortex were able to show significant reductions in tinnitus distress [26,27].

In addition to an rTMS individualization based on behavioral responses, the current investigation has striven for a novel attempt in rTMS individualization with objective measures. By the use of TMS-EEG combinatory measurements, we wanted to overcome the limitations of subjective tinnitus loudness ratings and identify personalized rTMS protocols via sham-superior increases in alpha respectively decreases in gamma frequency band power in at least two neighboring EEG electrodes on both test session. The utilization of especially these frequency bands as an electrophysiological response indicator is based on current neurophysiological models. In tinnitus pathological diminished alpha band activity was observed in contrast to healthy controls [57,58], whereby during states of brief tinnitus suppression either after sound stimulation [91] or after rTMS [46] increases in alpha power were detected. While gamma band activity is assumed to underly the actual tinnitus perception [57,59] by associations with tinnitus loudness [54,60]. Interestingly, we were able to identify a substantially higher number of EEG responders than behavioral responders. For a total number of 20 participants, it was possible to detect an alpha response for at least one rTMS protocol, whereas the entire sample featured responses in the gamma frequency band. Noteworthy, the quantity of alpha or gamma responders was almost twice as much as for behavioral responders. In line with our behavioral observations, a left 10 Hz stimulation resulted in the highest quantity of alpha and gamma responders. It has been shown that this type of stimulation can modulate alpha oscillations [92] and that alpha and gamma oscillations show a strong interplay not only in tinnitus [52,93].

Since neurophysiological investigations were able to demonstrate relationships of the alpha and gamma band with tinnitus loudness [46,54,60,91], we addressed the question if a response in the alpha or gamma band is associated with a behavioral response and vice versa. Our analysis did not reveal any relationship of EEG and behavioral responders, hampering the interpretability of our alpha and gamma findings as relevant for tinnitus loudness decreases in the current sample. A potential explanation for this lack of association could derive from the possibility, that rTMS-specific modulations/ neuroplastic consequences as measured with EEG simply do not instantly translate into behavioral responses. Consequently, longer periods of stimulation with an individualized rTMS protocol based on patients’ EEG response (protocol with strongest alpha increase/ gamma decrease) for example within the course of a daily treatment, might accumulate over time into behavioral effects and could tackle the deficiency of effectiveness in current rTMS treatment trials for tinnitus.

Future studies should strive for a systematic implementation of test session approaches and investigate the therapeutic consequences of personalized rTMS in more detail. The present experiment underscores the feasibility of rTMS individualization via behavioral or electrophysiological responses. The results and methodology described in this sample of tinnitus patients might encourage neuromodulation attempts in other pathologies to personalize rTMS in order to account for inter-subject variability in rTMS response.

## 5. Conclusion

The aim of this investigation was to examine the feasibility of rTMS individualization in tinnitus. This involved the identification of responders to specific rTMS protocols and the assessment of test-retest reliability of rTMS effects in tinnitus suppression and ongoing brain activity. We were able to demonstrate the feasibility of rTMS individualization by using test sessions with different rTMS protocols. Responses to specific protocols based on electrophysiological signatures could be identified in all patients, responses based on behavioral effects in the majority of patients. In contrast test-retest reliability as assessed with correlation analyses for the various rTMS protocols were rather low both for behavioral and electrophysiological effects.

Taken together these findings are highly encouraging for efforts to enhance the efficacy of rTMS by personalizing stimulation protocols.

## Data Availability

The datasets generated for this study are available on request to the corresponding author.

## 6. Conflict of Interest

The authors declare that they have no conflict of interest associated with this publication and there has been no significant financial support that could have influenced the outcomes. Author TH received a one-time travel cost coverage by Nexstim Plc. for an oral presentation.

## 7. Author Contributions

MS, BL and SS designed the study. MS and SS carried out the experiment. TH and MA recruited eligible participants and conducted the magnetic resonance imaging. SS analyzed the data and wrote the main manuscript under the supervision of MS. Authors BL, TH, MA and GV contributed to and reviewed the manuscript.

## 8. Funding

The project was conducted as part of the European School for Interdisciplinary Tinnitus Research (ESIT) and was partially funded by the European Union’s Horizon 2020 Marie Sklodowska-Curie Actions (grant agreement number 722046, ESIT project) and the European Union’s Horizon 2020 Framework Programme (grant agreement number 848261, UNITI project).

## 9. Acknowledgments

We want to thank Nexstim Plc. for providing the electric-field guided neuronavigation rTMS system to perform this feasibility trial.

## Notes

### Competing Interest Statement

The authors have declared no competing interest.

### Author Declarations

The study was approved by the ethics committee of the University of Regensburg, Germany (ethical approval number: 17-820-101)

