## Supplementary figures and images for "Heading for personalized rTMS in tinnitus: reliability of individualized stimulation protocols in behavioral and electrophysiological responses" (from another version of this preprint: version 1, 2021-01-13)

### Supplemental Figure S1

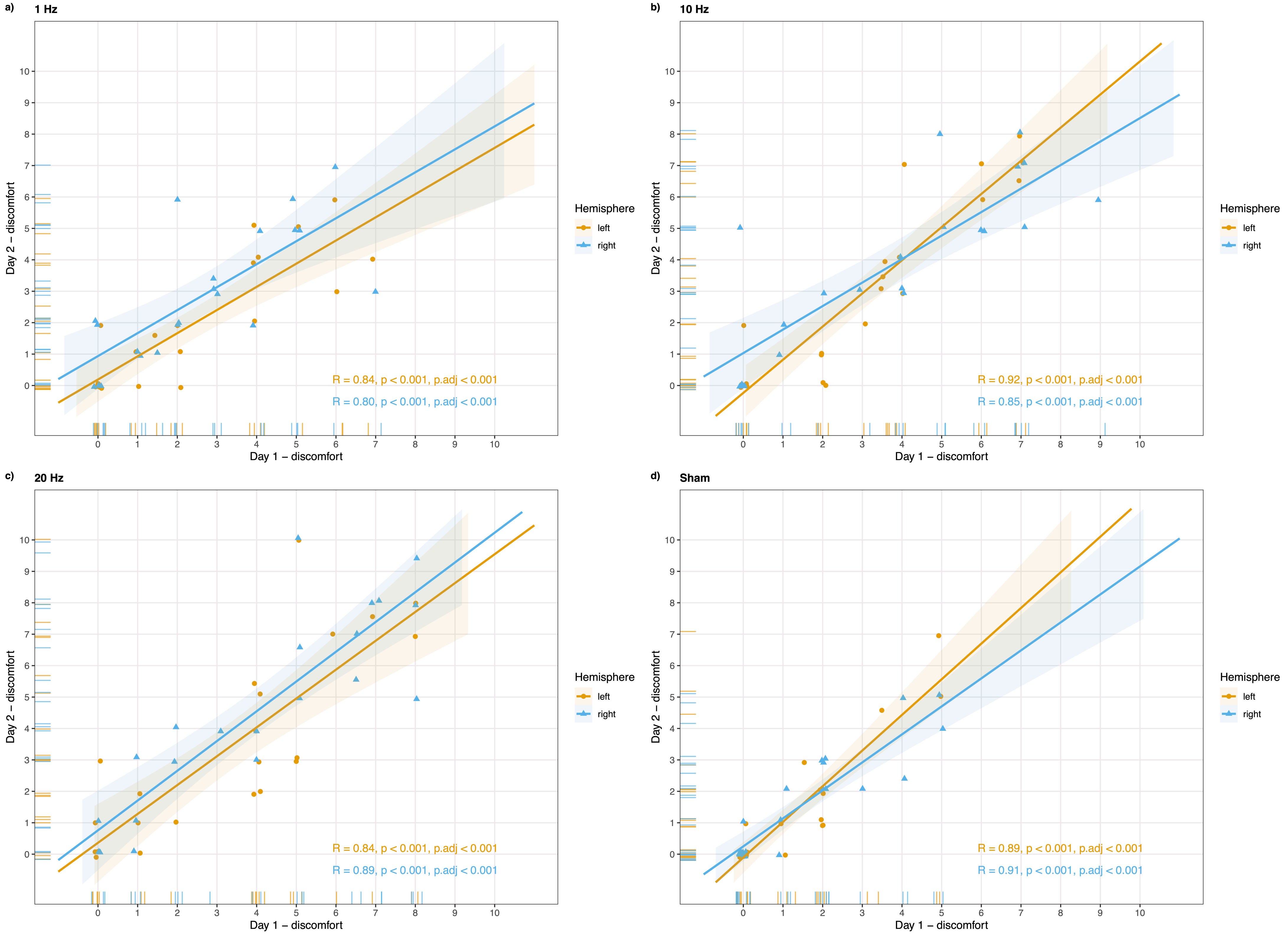
